# Cohort profile: the Johns Hopkins COVID Long Study (JHCLS), a United States Nationwide Prospective Cohort Study

**DOI:** 10.1101/2024.04.25.24306387

**Authors:** Eryka Wentz, Zhanmo Ni, Karine Yenokyan, Candelaria Vergara, Jessica Pahwa, Thea Kammerling, Pu Xiao, Priya Duggal, Bryan Lau, Shruti H. Mehta

## Abstract

**Purpose:** COVID-19 disease continues to affect millions of individuals worldwide, both in the short and long term. The post-acute complications of SARS-CoV-2 infection, referred to as long COVID, result in diverse symptoms affecting multiple organ systems. Little is known regarding how the symptoms associated with long COVID progress and resolve over time. The Johns Hopkins COVID Long Study aims to prospectively examine the short- and long-term consequences of COVID-19 disease in individuals both with and without a history of SARS-CoV-2 infection using self-reported data collected in an online survey.

**Participants:** Sixteen thousand, seven hundred sixty-four adults with a history of SARS-CoV-2 infection and 799 adults without a history of SARS-CoV-2 infection who completed an online baseline survey.

**Findings to date:** This cohort profile describes the baseline characteristics of the Johns Hopkins COVID Long Study. Among 16,764 participants with a history of SARS-CoV-2 infection and defined long COVID status, 75% reported a good or excellent health status prior to infection, 99% reported experiencing at least one COVID-19 symptom during the acute phase of infection, 9.9% reported a hospitalization, and 63% were defined as having long COVID using the WHO definition.

**Future plans:** Analysis of longitudinal data will be used to investigate the progression and resolution of long COVID symptoms over time.

## INTRODUCTION

Since its emergence in 2019, COVID-19 has greatly affected the health and well-being of millions of people worldwide.(1,2) Both acute and persistent post-infection complications have been reported by patients(3) and COVID-19 is now recognized as a multi-organ disease.(4) The World Health Organization (WHO) defines persistent post-infection complications, referred to as long COVID, as new or continuing symptoms three months after initial illness that last at least two months and cannot be explained otherwise.(5) Despite recent studies suggesting that long COVID may occur in 10-55% of individuals exposed to severe acute respiratory syndrome coronavirus 2 (SARS-CoV-2),(6–8) the exact incidence remains unknown. There is also uncertainty in the pathophysiology and symptomatology of long COVID.(9,10) With the elevated burden of COVID-19 worldwide, it is important to understand the full range of symptoms and long-term outcomes.(2) Moreover, the large number of individuals requiring continued medical care will pose an economic burden on our health care system.(10)

Similar to other infections, SARS-CoV-2 is associated with post-acute infection syndromes resulting in a variety of symptoms.(11) Some of the core symptoms associated with long COVID are common to other post-acute infection syndromes as well, including but not limited to fatigue, exertion intolerance, and neurocognitive impairment.(11) Despite our general knowledge of the occurrence of post-acute infection syndromes, it is largely understudied. Cohort studies composed of those who have had SARS-CoV-2 and those who have not (i.e., control population) are critical to understanding the gaps in our knowledge of long COVID and post-acute infections in general.

The presentation of those with long COVID is often marked with multiple diverse symptoms affecting multiple organs; each individual may have their own unique clinical presentation.(12) Though age is a major risk factor in COVID-19 related mortality, and despite a preponderance of long COVID among those aged 40 - 60 years, long COVID is reported across the age spectrum.(13) Similarly, long COVID is reported by persons of all genders, race/ethnicities, and those with and without pre-existing comorbidities.(13,14) Hence, it is essential that research both identifies and characterizes the main clinical and epidemiological features associated with long COVID, including potential targets for intervention.

For these reasons, the Johns Hopkins COVID Long Study (JHCLS) was established to prospectively examine the short- and long-term consequences of COVID-19 over a 3-year follow-up period. The overall objectives of the JHCLS are to (1) characterize the spectrum of long-term sequelae of SARS-CoV-2 infection; (2) identify individuals at risk for long-term sequelae; and (3) characterize the physical and mental health disability associated with long COVID. To meet study objectives, the cohort includes participants with and without a history of SARS-CoV-2 infection. This cohort profile describes baseline demographic and clinical characteristics of United States (U.S.) participants enrolled in the JHCLS.

## COHORT DESCRIPTION

### Study design and participants

The JHCLS launched for participants with a self-reported history of SARS-CoV-2 infection on February 2, 2021, and expanded to include participants without a history of SARS-CoV-2 infection on March 2, 2022. All consenting participants are asked to complete a one-time, short online baseline survey with the option to remain anonymous. At the end of the baseline survey, participants are asked if they agree to be contacted for future COVID-19 studies, such as enrollment into longitudinal follow-up. If they respond yes, they are contacted by email or phone 3-6 months later with information about participating in longitudinal follow-up. If they subsequently consent to participate in longitudinal follow-up, they are emailed a follow-up survey every 3-6 months.

As of February 14, 2023, 20,319 participants with a self-reported history of SARS-CoV-2 infection and 1,041 participants without a history of SARS-CoV-2 infection consented to participate in the JHCLS (Figure 1). Of the 20,319 participants with a history of infection, 15,478 with a defined long COVID status (76%) completed the baseline survey and 11,924 (59%) consented to be contacted for future studies. Of these, 6,327 have enrolled in longitudinal follow-up and completed their first follow-up survey. Of the 1,041 participants without a history of infection, 799 (77%) completed the baseline survey and 501 (48%) consented to be contacted for future studies. Of these, 278 have enrolled in longitudinal follow-up and completed their first follow-up survey. At each round of follow-up, participants without a history of infection are asked if they have experienced COVID-19 symptoms or tested positive for SARS-CoV-2 since their last survey completion. If they respond yes, they are transferred to the survey for participants with a history of infection. As of February 14, 2023, 46 of the 278 participants (17%) who completed their first round of longitudinal follow-up have self-reported either a positive SARS-CoV-2 test or symptoms of COVID-19. The median survey completion time is 20 minutes for the baseline survey and 24 minutes for the first follow-up survey.

**Figure 1.**
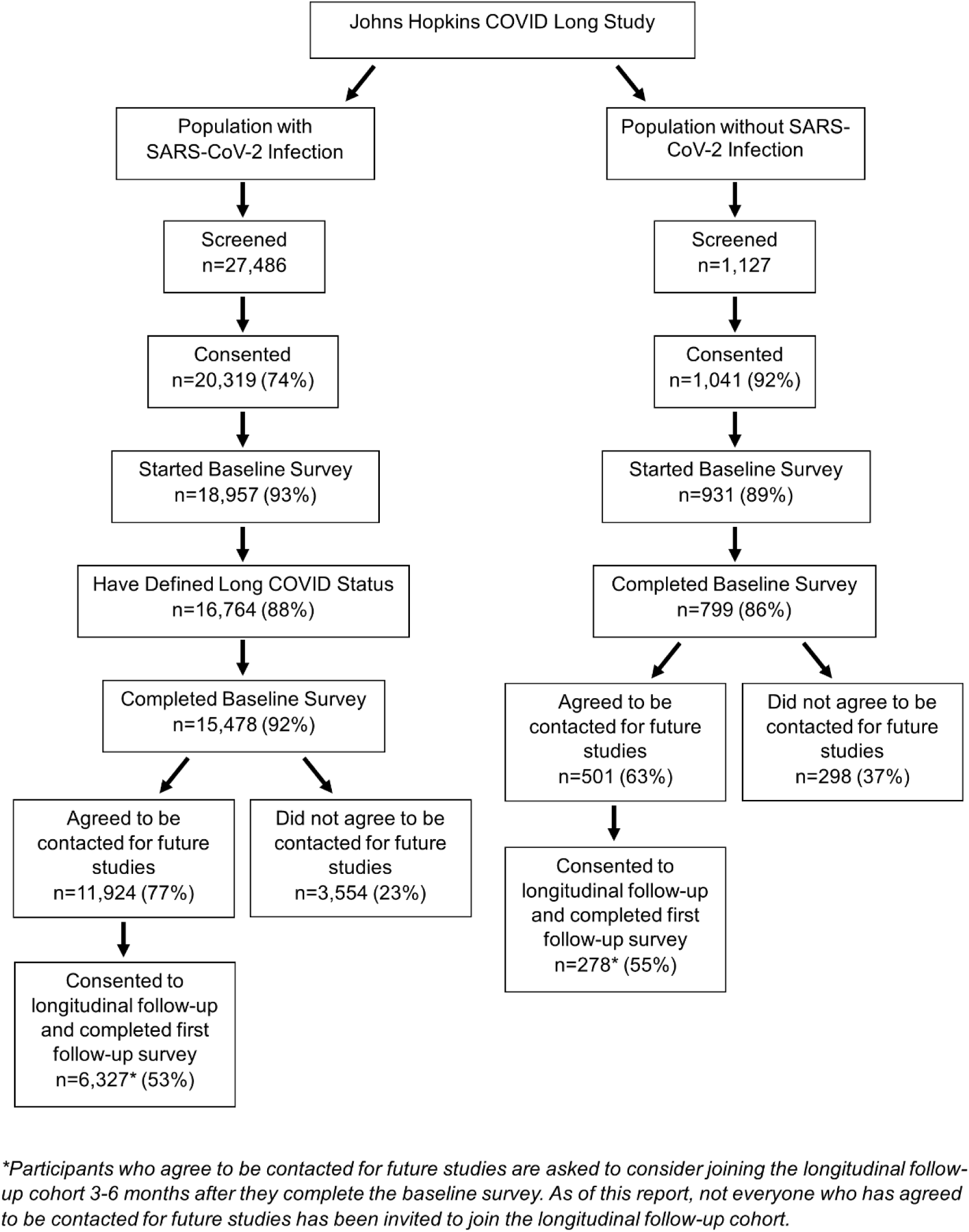
Flow diagram showing United States recruitment into the Johns Hopkins COVID Long Study

### Recruitment

Participants are recruited into the JHCLS using several mechanisms: social media posts, Facebook ad campaigns, direct messaging (e.g., emails to health departments), word of mouth, and participation in a recruitment registry. For social media recruitment, researchers utilize study-owned and operated Instagram, Facebook, and Twitter accounts. In addition, the team partnered with the Audience Development Team at the Johns Hopkins Bloomberg School of Public Health (BSPH) Communications Department to develop targeted Facebook ad campaigns. The study ran three Facebook ad campaigns, each targeting a neighborhood in the U.S. with high SARS-CoV-2 case counts. Two campaigns ran in April 2021, the first targeting neighborhoods in Detroit, Michigan, and the second in Fayetteville and Hope Mills, North Carolina, and South Fulton and Alpharetta, Georgia. The final campaign ran in July 2021 and targeted neighborhoods in Houston and San Antonio, Texas, Miami and Jacksonville, Florida, and Los Angeles, California.

The study team also partners with the Johns Hopkins Opportunities for Participant Engagement (HOPE) Registry. The HOPE Registry (http://johnshopkinshope.org/) is a recruitment registry designed to connect individuals with teams conducting COVID-19 research studies at Johns Hopkins University. The JHCLS was officially enrolled into the HOPE Registry in April 2021.

### Participant eligibility

The JHCLS was approved and determined to be exempt by the Institutional Review Board (IRB) at the BSPH on January 8, 2021.

To be eligible to participate in the study, participants must be at least 18 years of age. Additionally, to be eligible to complete the survey for participants with a history of SARS-CoV-2 infection, participants must self-report at least one positive SARS-CoV-2 test or symptoms of COVID-19. At the start of the baseline survey, eligible participants are provided with a short, informed consent script that details the purpose of the study and provides details on participation. In order to protect the confidentiality of participants, participants are assigned a unique study identifier number and data are collected anonymously.

### Study procedures

The JHCLS baseline survey is self-administered and collects data across nine domains: SARS-CoV-2 testing and COVID-19 symptoms, vaccines and SARS-CoV-2 re-infection, COVID-19 treatments and hospitalizations, pre-existing comorbidities, physical limitations and exercise, sleep quality, mental fatigue, anxiety, and demographics (Supplementary Table 1). Data from these same domains are collected during longitudinal follow-up as well. All data are collected in REDCap, a HIPAA-compliant, secure web application designed to build and manage online surveys and databases.(15,16) Most survey questions were adapted from validated measures and assessments. However, certain questions were self-designed for the purpose of meeting study objectives. All self-designed questionnaires are available on the National Institute of Environmental Health Sciences Disaster Research Response (DR2) Resources Portal (https://tools.niehs.nih.gov/dr2/index.cfm/resource/24278).

### SARS-CoV-2 Testing and COVID-19 Symptoms

To obtain data on COVID-19 history, diagnosis, and symptoms, researchers self-designed questions to assess overall health status prior to initial COVID-19 illness, history of SARS-CoV-2 testing and results, initial symptom onset date, symptoms experienced during the acute phase of COVID-19, new/continuing COVID-19 symptoms, impact of each reported symptom on daily activities, and self-reported recovery from COVID-19 illness. Participants without a history of infection are asked about health status prior to the COVID-19 pandemic, symptoms experienced in reference to overall general health, and self-reported recovery from the effects of the COVID-19 pandemic.

### Vaccines and SARS-CoV-2 Re-infection

To collect data on vaccination and SARS-CoV-2 re-infection, researchers self-designed questions related to flu vaccination uptake, COVID-19 vaccination uptake, SARS-CoV-2 antibody testing, participation in COVID-19 treatment trials, SARS-CoV-2 re-infection, and self-reported comparison of COVID-19 symptoms experienced during the first re-infection compared to initial illness. Participants without a history of infection are asked questions related to flu and COVID-19 vaccination uptake.

### Treatments and Hospitalizations

Researchers self-designed questions to obtain data on medications used to treat initial COVID-19 illness, medications used to treat new/continuing COVID-19 symptoms, COVID-19 related hospitalizations, health care utilization, and health seeking behavior to treat symptoms. In this section, participants without a history of infection are asked about overall health care utilization.

### Pre-existing Comorbidities

In order to obtain data on pre-existing health status and comorbidities, researchers self-designed questions to capture current health status, pre-existing health conditions, cancer diagnosis, height, weight, current stress level, and stress level prior to the COVID-19 pandemic.

### Physical Limitations & Exercise

To assess physical limitations and exercise, researchers utilized questions adapted from the Baltimore Longitudinal Study of Aging and the Godin-Shephard Leisure-Time Physical Activity Questionnaire.

The Baltimore Longitudinal Study of Aging (BLSA) is a longitudinal study of healthy adults with the aim of understanding how adults adjust to the aging process, including adjustments in physical activity.(17,18) During the baseline survey, participants are asked questions to assess difficulty and level of difficulty in the following domains: mobility (walking a quarter mile/one mile and going up 10 steps/20 steps) and instrumental activities of daily living (IADL) (light and heavy housework). If a participant reports experiencing difficulty, they are asked to report the level of difficulty (a little, some, a lot, or unable to do); conversely, if they do not report difficulty, they are asked to report the level of ease (very easy, somewhat easy, or not so easy).(17,18) Participants who report difficulty are also asked if they experienced the difficulty prior to their COVID-19 illness or the COVID-19 pandemic. For mobility, if a participant reports any level of difficulty walking a quarter of a mile, they are considered to have a mobility disability. For IADL, if a participant reports any level of difficulty with light housework, they are considered to have an IADL disability.

The Godin-Shephard Leisure-Time Physical Activity Questionnaire (GSLTPAQ) was validated for use in healthy adults by measuring the correlation between objective measures of physical condition, maximum oxygen intake during exercise (V02 max) and body fat percentile, and subjective measures of total leisure time physical activity.(19–21) The questionnaire was found to have a test-retest reliability of 0.94, 0.46, and 0.48 for strenuous, moderate, and light intensity exercise, respectively, with the highest correlation shown between V02 max and strenuous intensity exercise (Pearson’s r = 0.38) and body fat percentile and strenuous intensity exercise (Pearson’s r = 0.21).(20)

During the baseline survey, participants are asked to report the number of times on average they participate in mild, moderate, and strenuous intensity exercise for longer than 15 minutes during a typical week. The number of times per week is multiplied by the corresponding Metabolic Equivalent of Task (MET) factor (3, 5, and 9 for mild, moderate, and strenuous intensity exercise, respectively) and summed for a total leisure activity score.(19,20) A score of ≥24 indicates an active lifestyle, a score of 14-23 indicates a moderately active lifestyle, and a score of <14 indicates an insufficiently active/sedentary lifestyle.(19) Participants with a history of infection are asked to report the number of times they exercised in each category before and after their COVID-19 illness; participants without a history of infection are asked in reference to before and after the COVID-19 pandemic.

### Sleep Quality

Sleep quality is assessed using questions adapted from the AIDS Linked to the IntraVenous Experience (ALIVE) Study and the Idiopathic Hypersomnia Severity Scale. The ALIVE study is a prospective cohort study designed to characterize the incidence and natural history of HIV infection among injection drug users in Baltimore, MD.(22) Participants are asked how often they experience a list of five items related to sleep quality over the past four weeks. For each item, participants respond based on the following scale: 1 (all of the time), 2 (most of the time), 3 (a good bit of the time), 4 (some of the time), 5 (a little bit of the time), or 6 (none of the time).

The Idiopathic Hypersomnia Severity Scale (IHHS) was validated for use in patients experiencing three major symptoms of idiopathic hypersomnia: excessive daytime sleepiness, prolonged nighttime sleep, and sleep inertia, and was found to have high internal consistency (Cronbach α =.89) and good content validity.(23,24) The scale consists of 14 items and each item is scored separately and then summed together for a total score ranging from 0-50. Higher scores represent more severe/frequent symptoms of idiopathic hypersomnia.(23,24) For the purpose of the JHCLS, researchers utilized four questions from the IHHS for a range of scores from 0-14.

### Mental Fatigue

Mental fatigue is assessed using the Wood Mental Fatigue Inventory (WMFI). The WMFI has been validated for use in patients with Myalgic Encephalomyelitis/Chronic Fatigue Syndrome (ME/CFS) and was found to have high internal consistency (Cronbach α =.93) and good test-retest reliability (Pearson’s r = 0.887).(25) Participants are asked how much they have been bothered by a list of nine items over the past two weeks. Each item is scored on the following scale: 0 (not at all), 1 (a little), 2 (somewhat), 3 (quite a lot), or 4 (very much). At the end of the assessment, the scores are summed together for a range of 0-36. Higher scores indicate greater levels of mental fatigue.(25)

### Anxiety

To assess anxiety, researchers utilize the Generalized Anxiety Disorder-7 (GAD-7). The GAD-7 has been validated for use in the general population and was found to have both high internal consistency (Cronbach α =.92) and good criterion, construct, factorial, and procedural validity.(26) Participants are asked how often they have been bothered by a list of seven items over the past two weeks. For each item, participants are scored based on the following scale: 0 (not at all), 1 (several days), 2 (more than half the days), or 3 (nearly every day). At the end of the assessment, the scores are summed together for a range of 0-21. A score of 0-4 indicates no anxiety disorder, a score of 5-9 indicates a mild anxiety disorder, a score of 10-14 indicates a moderate anxiety disorder, and a score of ≥15 indicates a severe anxiety disorder.(26)

### Demographics

Researchers self-designed survey questions to obtain demographic information, including gender, race/ethnicity, country of residence, year of birth, educational attainment, work activities prior to the COVID-19 pandemic, primary occupation, total household income, and total number of dependents.

### Patient and public involvement

There was no patient or public involvement in the design, conduct, reporting, or dissemination plans of our research. However, patient feedback is routinely discussed and considered. Specifically, patients are encouraged to reach out to study team members with suggestions on ways to improve the survey and the survey has been adjusted several times based on patient suggestions. In addition, study findings are regularly disseminated to patients via quarterly study newsletters posted on the study website (www.covid-long.com).

### Baseline characteristics of JHCLS participants

Among 16,764 participants with a history of SARS-CoV-2 infection and defined long COVID status, the median age was 43 years, 84% were female, 88% self-reported white race, and 8.0% self-reported Hispanic/Latino ethnicity (Table 1). In terms of socioeconomic status, 70% of participants self-reported a bachelor’s degree or higher and 72% self-reported an annual household income of greater than or equal to $50,000. A diverse array of self-reported pre-existing comorbid conditions were reported, including hypertension (15%), depression/anxiety/other mental health conditions (35%), asthma/reactive airway disease/chronic lung disease (16%), and autoimmune disorders (9.6%) (Table 2). In addition, the majority of participants (65%) were classified as overweight/obese based on a calculated BMI of ≥25.

**Table 1.**
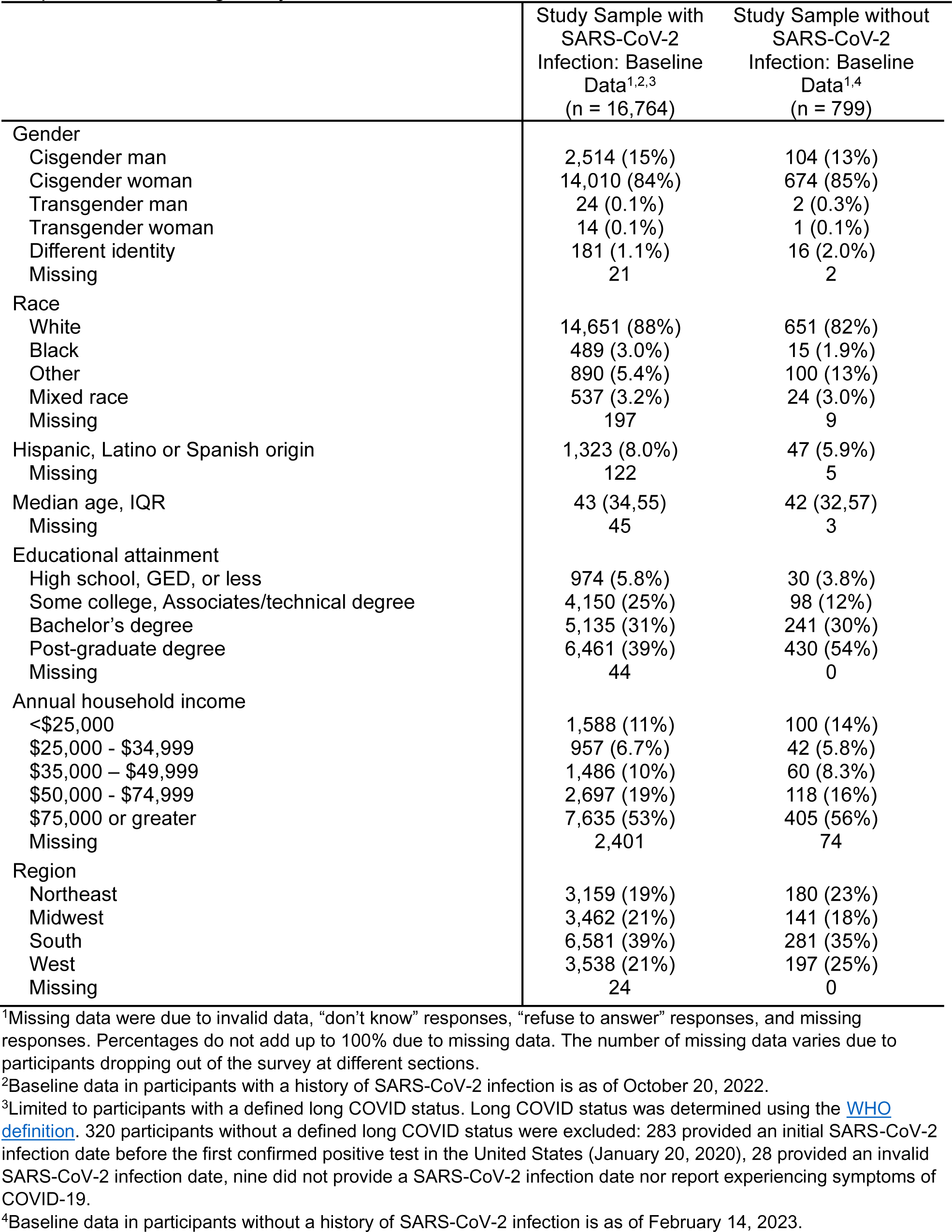
Baseline demographic characteristics of United States participants in the Johns Hopkins COVID Long Study.

**Table 2.** Baseline clinical characteristics of United States participants in the Johns Hopkins COVID Long Study.

|  | Study Sample with<br>SARS-CoV-2<br>Infection: Baseline<br>Data<sup>1,2</sup><br>(n = 16,764) | Study Sample without<br>SARS-CoV-2<br>Infection: Baseline<br>Data<sup>1,3</sup><br>(n = 799) |
| --- | --- | --- |
| Body mass index (kg/m<sup>2</sup>) |  |  |
| Underweight (<18.5) | 285 (1.8%) | 13 (1.6%) |
| Normal weight (18.5 – 24.9) | 5,517 (34%) | 342 (43%) |
| Overweight (25 – 29.9) | 4,643 (29%) | 216 (27%) |
| Obese (30 and above) | 5,827 (36%) | 225 (28%) |
| Missing | 492 | 3 |
| Comorbid conditions |  |  |
| Diabetes | 716 (4.4%) | 32 (4.0%) |
| Cardiovascular disease/congestive heart failure | 379 (2.3%) | 24 (3.0%) |
| Hypertension | 2,526 (15%) | 105 (13%) |
| Chronic kidney disease | 138 (0.8%) | 6 (0.8%) |
| Cancer | 392 (2.4%) | 17 (2.1%) |
| Asthma/reactive airway disease/chronic lung disease | 2,684 (16%) | 99 (12%) |
| Overweight/obese | 4,853 (30%) | 187 (23%) |
| Autoimmune disorder | 1,568 (9.6%) | 65 (8.1%) |
| Stroke | 116 (0.7%) | 9 (1.1%) |
| Depression/anxiety/other mental health condition | 5,730 (35%) | 286 (36%) |
| Missing | 388 | 0 |
| Self-rated health status prior to COVID-19<sup>4</sup> |  |  |
| Excellent | 6,297 (38%) | 283 (35%) |
| Very good | 6,192 (37%) | 321 (40%) |
| Good | 2,900 (17%) | 135 (17%) |
| Fair | 1,242 (7.4%) | 53 (6.6%) |
| Poor | 126 (0.8%) | 7 (0.9%) |
| Missing | 7 | 0 |
| Vaccination status at time of enrollment |  |  |
| None = 0 | 6,414 (39%) | 22 (2.8%) |
| Partial vaccination | 805 (4.9%) | 4 (0.5%) |
| Complete first series | 4,006 (24%) | 93 (12%) |
| ≥1 Booster | 5,208 (32%) | 679 (85%) |
| Missing | 331 | 1 |
| Timing of initial SARS-CoV-2 infection |  |  |
| January - June 2020 | 3,804 (23%) | N/A |
| July - December 2020 | 5,484 (33%) | N/A |
| January - June 2021 | 2,216 (13%) | N/A |
| July - December 2021 | 2,629 (16%) | N/A |
| ≥January 2022 | 2,631 (16%) | N/A |
| Missing | 0 | N/A |
| Time between initial infection and survey completion in days (median (IQR)) | 173 (70,382) | N/A |
| Missing | 0 | N/A |
| Symptom status at initial COVID-19 illness |  |  |
| Symptomatic | 16,588 (99%) | N/A |
| Asymptomatic | 175 (1.0%) | N/A |
| Missing | 1 | N/A |
| Presenting symptoms at initial COVID-19 illness |  |  |

|  | Study Sample with<br>SARS-CoV-2<br>Infection: Baseline<br>Data<sup>1,2</sup><br>(n = 16,764) | Study Sample without<br>SARS-CoV-2<br>Infection: Baseline<br>Data<sup>1,3</sup><br>(n = 799) |
| --- | --- | --- |
| Cardiopulmonary | 15,140 (90%) | N/A |
| Neuropsychiatric | 14,178 (85%) | N/A |
| Systemic | 14,949 (89%) | N/A |
| Gastrointestinal | 9,175 (55%) | N/A |
| Missing | 0 | N/A |
| Hospitalization status at initial COVID-19 illness |  |  |
| Not hospitalized | 14,839 (90%) | N/A |
| Hospitalized | 1,627 (9.9%) | N/A |
| Missing | 298 | N/A |
| Long COVID status at survey completion<sup>5</sup> |  |  |
| Has long COVID | 10,518 (63%) | N/A |
| Does not have long COVID | 1,246 (7.4%) | N/A |
| Cannot be determined<sup>6</sup> | 5,000 (30%) | N/A |

<sup>1</sup>Missing data were due to invalid data, “don’t know” responses, “refuse to answer” responses, and missing responses. Percentages do not add up to 100% due to missing data. The number of missing data varies due to participants dropping out of the survey at different sections.
<sup>2</sup>Baseline data in participants with a history of SARS-CoV-2 infection is as of October 20, 2022.
<sup>3</sup>Baseline data in participants without a history of SARS-CoV-2 infection is as of February 14, 2023.
<sup>4</sup>Participants with a history of SARS-CoV-2 infection were asked for self-rated health status prior to their COVID-19 illness. Participants without a history of SARS-CoV-2 infection were asked for self-rated health status prior to the COVID-19 pandemic.
<sup>5</sup>Limited to participants with a defined long COVID status. Long COVID status was determined using the WHO definition. 320 participants without a defined long COVID status were excluded: 283 provided an initial SARS-CoV-2 infection date before the first confirmed positive test in the United States (January 20, 2020), 28 provided an invalid SARS-CoV-2 infection date, nine did not provide a SARS-CoV-2 infection date nor report experiencing symptoms of COVID-19.
<sup>6</sup>Long COVID status could not be determined because fewer than 12 weeks existed between initial SARS-CoV-2 infection and survey completion.

Prior to COVID-19 illness, 75% of participants reported very good/excellent health status and 8.2% of participants reported fair/poor health status (Table 2). During the acute phase of COVID-19 illness, 99% of participants reported experiencing at least one symptom. Of those, 90% reported cardiopulmonary symptoms (e.g., new/worsening cough, shortness of breath, rapid heart rate), 89% reported systemic symptoms (e.g., fatigue, muscle weakness, fever), 85% reported neuropsychiatric symptoms (e.g., headache, dizziness, neuropathy), and 55% reported gastrointestinal symptoms (e.g., vomiting, diarrhea, lack of appetite). Overall, 9.9% of participants self-reported being hospitalized for their COVID-19 illness and 63% were defined as having long COVID based on the WHO definition.

At the time of study enrollment, 39% of participants with a history of infection reported not being vaccinated against SARS-CoV-2 compared to 56% who reported receiving at least a complete first vaccination series (Table 2). The median number of days between initial SARS-CoV-2 infection and study enrollment was 173 days. While most participants reported experiencing their initial SARS-CoV-2 infection in 2020 (56%), 29% reported being infected in 2021, and 16% reported being infected in 2022.

Similar characteristics were found in participants without a history of SARS-CoV-2 infection who completed the baseline survey. Among 799 participants, the median age was 42, 85% were female, 82% self-reported white race, and 5.9% self-reported Hispanic/Latino ethnicity (Table 1). A higher percentage of participants without a history of infection self-reported ‘other’ race (13% compared to 5.4%) which was largely due to a greater number self-reporting Asian/Pacific Islander/Native Hawaiian race. With regard to socioeconomic status, a higher percentage of participants without a history of infection reported a bachelor’s degree or higher (84% compared to 70%) and 72% reported an annual household income of $50,000 or more. Comparable pre-existing comorbid conditions were reported: hypertension (13%), depression/anxiety/other mental health conditions (36%), asthma/reactive airway disease/chronic lung disease (12%), and autoimmune disorders (8.1%) (Table 2). Based on calculated BMI, a slightly higher percentage of participants without a history of infection were classified as having a normal BMI (43% compared to 34%).

Of participants without a history of SARS-CoV-2 infection, 75% reported very good/excellent health status and 7.5% reported fair/poor health status prior to the COVID-19 pandemic (Table 2). At time of study enrollment, a higher percentage of participants without a history of infection reported at least a complete first vaccination series (97% compared to 56%). It is worth noting that enrollment for participants without a history of infection opened up in March 2022 when vaccinations were more widely available, likely accounting for this difference.

The demographic and clinical characteristics between those who agreed to be contacted for future studies and those who declined were comparable, with the exception of long COVID status (Supplementary Table 2). Unsurprisingly, more individuals who fully recovered declined continued participation in the study. However, the number of indeterminate individuals (too early to determine long COVID status) was similar.

## STRENGTHS AND LIMITATIONS

The JHCLS is a large, online, prospective cohort study of adults with representation in 53 U.S. states and territories. The baseline survey collects comprehensive clinical and behavioral data, including data related to COVID-19 diagnosis and treatment, health history, pre-existing health conditions, and physical, mental, and cognitive limitations, and utilizes several reliable, validated scales to assess outcomes and exploratory variables. Participants are given the option to complete a one-time, anonymous online survey or to consent to longitudinal follow-up at predefined time intervals (every 3-6 months). In addition, the overall participant burden is minimal.

A major strength of the JHCLS is that a positive SARS-CoV-2 test is not required to be eligible to participate. We recognize that testing is often limited or inaccessible, and thus require either a self-reported positive test or symptoms of COVID-19. In addition, our survey collects data on a wide range of organ systems using several different validated measures. Despite early studies focusing primarily on the respiratory symptoms associated with initial COVID-19 illness (e.g., shortness of breath), we appreciate that the SARS-CoV-2 virus may have notable effects on other organ systems following the acute period of infection as well.

Another strength of the JHCLS is the inclusion of participants without a history of SARS-CoV-2 infection which provides a natural control group, while also allowing for the determination of the incidence of long COVID among those who report a SARS-CoV-2 infection during follow-up. Importantly, both samples are comparable in terms of sociodemographic variables and pre-existing health conditions. We also recognize that many of the heterogeneous symptoms reported as long COVID may reflect all of us collectively living through a pandemic (i.e., anxiety, depression). Thus, it is important that we compare those with and without infection to evaluate some of these outcomes during the same time frame (versus retrospective or historical controls).

In addition, there are few longitudinal studies focused on post-acute outcomes of COVID-19. Longitudinal studies provide an opportunity to evaluate change over time in exposures and outcomes. The longitudinal collection of data on new/continuing COVID-19 symptoms at each time point during follow-up will allow for evaluation of resolution and persistence of symptoms over time, as well as the impact of re-infection, vaccination, and other health changes. To date, just under 7,000 participants have consented to participate in longitudinal follow-up and have completed their first follow-up survey.

The JHCLS also has a few limitations. The reliance on self-reported clinical data, including self-reported SARS-CoV-2 tests, may result in recall and measurement bias. Although a confirmed SARS-CoV-2 test would be preferable, we recognize that tests were not available to everyone and that restriction to only those with a confirmed test would introduce selection bias. A second limitation is the possibility of selection bias due to the fact that the survey must be completed using a smart device or computer with internet access. This may preclude participants from lower socioeconomic statuses from participating. There is also a risk that findings from the JHCLS are not generalizable as the majority of participants self-reported white race, female gender, and are from a higher socioeconomic status. To address this, we plan to do stratified-specific analyses that may be better representative of individuals within that same stratum. However, whether a study is representative or not depends not on demographics but on potential effect measure modifiers that may or may not include demographics.(27) Additionally, results that may not necessarily be generalizable in the effect estimate may still be generalizable in the direction of effect (e.g., protective or increased risk) of an exposure on outcome.(27)

Additionally, many individuals enroll many months after their acute infection when they already have long COVID. A potential selection bias would include increased likelihood of participation among those with more severe long COVID. However, it is important to note that the JHCLS has a subset of individuals (n = 2,020) who enrolled during their acute infection (within four weeks of infection). The high percentage of participants in our study with long COVID (63%) also likely reflects a selection bias on those willing to participate in COVID-19 research. However, those with and without a history of SARS-CoV-2 infection are similar in their demographic characteristics (Table 1). Another limitation is the possibility of recall bias, especially among participants with a history of COVID-19 illness experiencing mental fatigue and/or other cognitive limitations at the time of survey completion.

A final limitation is the use of non-validated instruments to collect COVID-19 related data, including COVID-19 diagnosis, treatments, and symptoms. We were limited by the unavailability of validated instruments to capture these domains at the time of study initiation. However, when available, we used validated instruments that targeted several specific domains (e.g., anxiety, mental fatigue, etc.), and when unavailable, we drew upon experience and validated instruments developed for other infectious diseases to develop questions used by our group and others across multiple COVID-19 studies.

### Future plans

Moving forward, the JHCLS will continue to enroll additional participants with and without a history of SARS-CoV-2 infection and collect data from the baseline and longitudinal surveys. The study team is in the initial stages of analyzing the longitudinal data collected thus far, focusing on the progression and resolution of long COVID symptoms over time. In addition, the study team is planning a cluster analysis of both initial and new/continuing COVID-19 symptoms to help address the broad WHO definition of long COVID. We plan to do this by bringing together the rich symptom data we have in our study with data on the impact each reported symptom has on daily functioning. In the future, the study team may apply for funding to answer additional research questions and to continue following the longitudinal participants for a longer period of time.

The JHCLS has the potential to impact both our overall understanding of long COVID and our ability to identify subgroups of individuals for targeted interventions. We can also capture real-time changes by SARS-CoV-2 variants (based on calendar time), location (using geospatial data), birth/age cohorts, and/or vaccine data.

## COLLABORATION

The JHCLS invites researchers to contact the corresponding author for collaboration opportunities.

## Data Availability

There was no data produced in the present study.

## Acknowledgements

We would like to express our deepest appreciation to our participants for their dedication, unwavering commitment, and vulnerability in sharing their stories with us. We would also like to thank the REDCap team at the Johns Hopkins Bloomberg School of Public Health for their continued assistance and guidance. This endeavor would not have been possible without their help. Lastly, we’d like to acknowledge and thank our student researchers.

## Author contributions

B. Lau, P. Duggal, and S.H. Mehta conceived the original study concept and design and act as Co-Principal Investigators. They take responsibility for the integrity of the data. B. Lau, P. Duggal, S.H. Mehta, and E. Wentz were responsible for the acquisition of the data. E. Wentz prepared the first draft of this manuscript, under the supervision of B. Lau, P. Duggal, and S.H. Mehta. E. Wentz, Z. Ni, K. Yenokyan, C. Vergara, J. Pahwa, T. Kammerling, P. Xiao, P. Duggal, B. Lau, and S.H. Mehta were involved in reviewing the manuscript and contributing to critical revisions. Administrative and technical support was provided by E. Wentz, C. Vergara, Z. Ni, J. Pahwa, T. Kammerling, and P. Xiao.

## Patient consent for publication

Not applicable.

## Ethics approval

The study protocol was approved and exempted by the Institutional Review Board at the Johns Hopkins Bloomberg School of Public Health (IRB00014874) on January 8, 2021.

## Data sharing statement

Not applicable.

## REFERENCES

1. Bourmistrova NW, Solomon T, Braude P, Strawbridge R, Carter B. Long-term effects of COVID-19 on mental health: A systematic review. J Affect Disord. 2022 Feb;299:118–25.

2. Nalbandian A, Sehgal K, Gupta A, Madhavan MV, McGroder C, Stevens JS, et al. Post-acute COVID-19 syndrome. Nat Med. 2021 Apr;27(4):601–15.

3. Raman B, Bluemke DA, Lüscher TF, Neubauer S. Long COVID: post-acute sequelae of COVID-19 with a cardiovascular focus. Eur Heart J. 2022 Mar 14;43(11):1157–72.

4. Montani D, Savale L, Noel N, Meyrignac O, Colle R, Gasnier M, et al. Post-acute COVID-19 syndrome. Eur Respir Rev. 2022 Mar 31;31(163):210185.

5. Soriano JB, Murthy S, Marshall JC, Relan P, Diaz JV. A clinical case definition of post-COVID-19 condition by a Delphi consensus. Lancet Infect Dis. 2022 Apr;22(4):e102–7.

6. Perlis RH, Santillana M, Ognyanova K, Safarpour A, Lunz Trujillo K, Simonson MD, et al. Prevalence and Correlates of Long COVID Symptoms Among US Adults. JAMA Netw Open. 2022 Oct 27;5(10):e2238804.

7. Van Kessel SAM, Olde Hartman TC, Lucassen PLBJ, Van Jaarsveld CHM. Post-acute and long-COVID-19 symptoms in patients with mild diseases: a systematic review. Fam Pract. 2022 Jan 19;39(1):159–67.

8. Domingo FR, Waddell LA, Cheung AM, Cooper CL, Belcourt VJ, Zuckermann AME, et al. Prevalence of long-term effects in individuals diagnosed with COVID-19: an updated living systematic review [Internet]. Epidemiology; 2021 Jun [cited 2023 Jun 28]. Available from: http://medrxiv.org/lookup/doi/10.1101/2021.06.03.21258317

9. Castanares-Zapatero D, Chalon P, Kohn L, Dauvrin M, Detollenaere J, Maertens De Noordhout C, et al. Pathophysiology and mechanism of long COVID: a comprehensive review. Ann Med. 2022 Dec 31;54(1):1473–87.

10. Rodriguez-Sanchez I, Rodriguez-Mañas L, Laosa O. Long COVID-19: The Need for an Interdisciplinary Approach. Clin Geriatr Med. 2022 Aug;38(3):533–44.

11. Choutka J, Jansari V, Hornig M, Iwasaki A. Unexplained post-acute infection syndromes. Nat Med. 2022 May;28(5):911–23.

12. Ramakrishnan RK, Kashour T, Hamid Q, Halwani R, Tleyjeh IM. Unraveling the Mystery Surrounding Post-Acute Sequelae of COVID-19. Front Immunol. 2021 Jun 30;12:686029.

13. Pavli A, Theodoridou M, Maltezou HC. Post-COVID Syndrome: Incidence, Clinical Spectrum, and Challenges for Primary Healthcare Professionals. Arch Med Res. 2021 Aug;52(6):575–81.

14. Karuna S, Gallardo-Cartagena JA, Theodore D, Hunidzarira P, Montenegro-Idrogo J, Hu J, et al. Post-COVID symptom profiles and duration in a global convalescent COVID-19 observational cohort: Correlations with demographics, medical history, acute COVID-19 severity and global region. J Glob Health. 2023 Jun 23;13:06020.

15. Harris PA, Taylor R, Thielke R, Payne J, Gonzalez N, Conde JG. Research electronic data capture (REDCap)—A metadata-driven methodology and workflow process for providing translational research informatics support. J Biomed Inform. 2009 Apr;42(2):377–81.

16. Harris PA, Taylor R, Minor BL, Elliott V, Fernandez M, O’Neal L, et al. The REDCap consortium: Building an international community of software platform partners. J Biomed Inform. 2019 Jul;95:103208.

17. Ferrucci L. The Baltimore Longitudinal Study of Aging (BLSA): a 50-year-long journey and plans for the future. J Gerontol A Biol Sci Med Sci. 2008 Dec;63(12):1416–9.

18. Stone JL, Norris AH. Activities and Attitudes of Participants in the Baltimore Longitudinal Study. J Gerontol. 1966 Oct 1;21(4):575–80.

19. Gaston Godin. The Godin--Shephard Leisure--Time Physical Activity Questionnaire. Health Fit J Can. 2011;4(1):18–22.

20. Godin G, Shephard RJ. A simple method to assess exercise behavior in the community. Can J Appl Sport Sci J Can Sci Appl Au Sport. 1985 Sep;10(3):141–6.

21. Amireault S, Godin G. The Godin-Shephard Leisure-Time Physical Activity Questionnaire: Validity Evidence Supporting its Use for Classifying Healthy Adults into Active and Insufficiently Active Categories. Percept Mot Skills. 2015 Apr;120(2):604–22.

22. Vlahov D, Anthony JC, Munoz A, Margolick J, Nelson KE, Celentano DD, et al. The ALIVE study, a longitudinal study of HIV-1 infection in intravenous drug users: description of methods and characteristics of participants. NIDA Res Monogr. 1991;109:75–100.

23. Rassu AL, Evangelista E, Barateau L, Chenini S, Lopez R, Jaussent I, et al. Idiopathic Hypersomnia Severity Scale to better quantify symptoms severity and their consequences in idiopathic hypersomnia. J Clin Sleep Med. 2022 Feb;18(2):617–29.

24. Dauvilliers Y, Evangelista E, Barateau L, Lopez R, Chenini S, Delbos C, et al. Measurement of symptoms in idiopathic hypersomnia: The Idiopathic Hypersomnia Severity Scale. Neurology. 2019 Apr 9;92(15):e1754–62.

25. Bentall RP, Wood GC, Marrinan T, Deans C, Edwards RHT. A brief mental fatigue questionnaire. Br J Clin Psychol. 1993 Sep;32(3):375–7.

26. Spitzer RL, Kroenke K, Williams JBW, Löwe B. A Brief Measure for Assessing Generalized Anxiety Disorder: The GAD-7. Arch Intern Med. 2006 May 22;166(10):1092.

27. Jacqueline E Rudolph, Yongqi Zhong, Priya Duggal, Shruti H Mehta, Bryan Lau. Defining representativeness of study samples in medical and population health research. BMJ Med. 2023 May 1;2(1):e000399.

