## Supplementary Table 1 for "Cohort profile: the Johns Hopkins COVID Long Study (JHCLS), a United States Nationwide Prospective Cohort Study"

| **Supplementary Table 1** Survey measures | | | | |
|---|---|---|---|---|
| **Domain** | **Questionnaire** | **Variable Description** | **Validation** | **Scoring** |
| SARS-CoV-2 Testing and COVID-19 Symptoms | Self-designed | General health before COVID-19 illness (population with history of SARS-CoV-2 infection) or COVID-19 pandemic (population without history of SARS-CoV-2 infection), SARS-CoV-2 testing (number of times, result, exposure, test type, month/year of first positive test), month/year of initial symptom onset, symptoms experienced during initial illness, new/continuing symptoms experienced after initial illness, impact of each symptom on daily activities, self-reported recovery compared to before COVID-19 illness (population with history of SARS-CoV-2 infection) or COVID-19 pandemic (population without history of SARS-CoV-2 infection), new physician diagnoses since COVID-19 illness (population with history of SARS-CoV-2 infection) or COVID-19 pandemic (population without history of SARS-CoV-2 infection) | N/A | Categorical responses |
| Vaccines and SARS-CoV-2 Re-infection | Self-designed | Flu vaccine uptake, COVID-19 vaccine uptake (number of doses, series type, month/year per dose), SARS-CoV-2 antibody testing (month/year of testing, result), COVID-19 treatment trials, COVID-19 re-infection (number of times, exposure, test type, month/year of each positive test, month/year of each symptom onset), comparison of first re-infection to initial infection | N/A | Categorical responses |
| COVID-19 Treatments and Hospitalizations | Self-designed | Treatments for COVID-19, treatments for new/continuing COVID-19 symptoms, hospitalizations (number of days, severity), health care utilization (pre-COVID-19 illness and current), health seeking behavior to treat symptoms | N/A | Categorical responses |
| Comorbidities | Self-designed | Self-reported current health status, pre-existing health conditions, cancer diagnosis (type, diagnosis timeframe, treatments), height, weight, current stress level, stress level before the COVID-19 pandemic | N/A | Categorical responses |

| **Supplementary Table 1** Continued | | | | |
|---|---|---|---|---|
| **Domain** | **Questionnaire** | **Variable Description** | **Validation** | **Scoring** |
| Limitations and Exercise | Baltimore Longitudinal Study of Aging(17,18) | Overall physical limitations before COVID-19 illness (population with history of SARS-CoV-2 infection) or COVID-19 pandemic (population without history of SARS-CoV-2 infection), difficulty walking a quarter of a mile/one mile, difficulty walking up 10 steps/20 steps, difficulty with performing light housework/heavy housework, difficulty level (if difficulty reported), level of ease (if no difficulty reported), indicator for incident/prevalent disability | N/A | Mobility disability: Any level of difficulty walking a quarter of a mile<br>Instrumental activities of daily living disability: Any level of difficulty with light housework |
| | Godin-Shephard Leisure-Time Physical Activity Questionnaire(19–21) | Number of times in a typical week doing strenuous, moderate, and mild intensity exercise for more than 15 minutes before and after COVID-19 illness (population with history of SARS-CoV-2 infection) or COVID-19 pandemic (population without history of SARS-CoV-2 infection) | Validated in population of healthy adults<br>Test-retest reliability: 0.94 for strenuous exercise, 0.46 for moderate exercise, and 0.48 for light exercise | 1) Multiply number of times per week per category by Metabolic Equivalent of Task factor (3 for light, 5 for moderate, 9 for strenuous)<br>2) Sum scores for total leisure time activity score<br>• >24: active lifestyle<br>• 14-23: moderately active lifestyle<br>• <14: sedentary lifestyle |
| Sleep Quality | AIDS Linked to the IntraVenous Experience (ALIVE) Study(22) | Total hours slept in typical 24-hour period, overall sleep quality during last four weeks, insomnia (difficulty falling/staying asleep) | N/A | Categorical responses |
| | Idiopathic Hypersomnia Severity Scale(23,24) | Four indicators of hypersomnia (assessment of sleep adequacy, difficulty waking up, length of time to feel fully functioning upon waking, struggling to stay awake during the day) | Validated in patients experiencing idiopathic hypersomnia<br>High internal consistency (Cronbach α=.89) and good content validity | 1) Each item is assigned a score (0-3 or 0-4)<br>2) Sum scores for a total of 0-14<br>Higher scores represent more severe/frequent symptoms of idiopathic hypersomnia |

| **Supplementary Table 1** Continued | | | | |
|---|---|---|---|---|
| **Domain** | **Questionnaire** | **Variable Description** | **Validation** | **Scoring** |
| Cognition | Wood Mental Fatigue Inventory (WMFI)(25) | Nine indicators of mental fatigue over last two weeks (confusion, mixed thoughts, poor concentration, difficulty with decision making, memory problems, issues taking things in, slow thoughts, muzzy head, issues finding words) | Validated in patients with ME/CFS<br>High internal consistency (Cronbach α=.93) and good test-retest reliability (Pearson's r = 0.887) | 1) Each item is assigned a score from 0-4<br>2) Sum scores for total of 0-36<br>Higher scores indicate greater levels of mental fatigue |
| Anxiety | Generalized Anxiety Disorder-7 (GAD-7)(26) | Seven indicators of anxiety over last two weeks (feeling anxious, not able to control worrying, worrying about different things, trouble relaxing, restlessness, irritability, feeling afraid) | Validated in general population<br>High internal consistency (Cronbach α=.92)<br>Good criterion, construct, factorial, and procedural validity | 1) Each item is assigned a score from 0-3<br>2) Sum scores for total of 0-21<br>• 0-4: no anxiety disorder<br>• 5-9: mild anxiety disorder<br>• 10-14: moderate anxiety disorder<br>• $\geq$15: severe anxiety disorder |
| Demographics | Self-designed | Work activities prior to the COVID-19 pandemic, primary occupation, household income in 2019, number of dependents | N/A | Categorical responses |
