## Supplementary Table 2 for "Cohort profile: the Johns Hopkins COVID Long Study (JHCLS), a United States Nationwide Prospective Cohort Study"

**Supplementary Table 2** Comparison of baseline demographic and clinical characteristics among participants with SARS-CoV-2 infection who agreed and declined to be contacted for future studies[1,2,3,4]

| | Agreed to be Contacted for Future Studies (n = 11,924) | Declined to be Contacted for Future Studies (n = 3,554) |
|---|---|---|
| Gender | | |
| Cisgender man | 1,829 (15%) | 510 (14%) |
| Cisgender woman | 9,934 (83%) | 2,992 (84%) |
| Transgender man | 15 (0.1%) | 6 (0.2%) |
| Transgender woman | 11 (0.1%) | 1 (<0.1%) |
| Different identity | 127 (1.1%) | 36 (1.0%) |
| Missing | 8 | 9 |
| Race | | |
| White | 10,538 (89%) | 3,067 (88%) |
| Black | 348 (2.9%) | 88 (2.5%) |
| Other | 563 (4.8%) | 213 (6.1%) |
| Mixed race | 384 (3.3%) | 117 (3.4%) |
| Missing | 91 | 69 |
| Hispanic, Latino or Spanish origin | 892 (7.5%) | 289 (8.2%) |
| Missing | 68 | 35 |
| Median age, IQR | 46 (36,57) | 42 (33,52) |
| Missing | 34 | 6 |
| Educational attainment | | |
| High school, GED, or less | 620 (5.2%) | 243 (6.9%) |
| Some college, Associates/technical degree | 3,009 (25%) | 823 (23%) |
| Bachelor’s degree | 3,601 (30%) | 1,116 (32%) |
| Post-graduate degree | 4,676 (39%) | 1,356 (38%) |
| Missing | 18 | 16 |
| Annual household income | | |
| <$25,000 | 1,163 (11%) | 394 (13%) |
| $25,000 - $34,999 | 712 (6.5%) | 227 (7.4%) |
| $35,000 – $49,999 | 1,125 (10%) | 328 (11%) |
| $50,000 - $74,999 | 2,084 (19%) | 576 (19%) |
| $75,000 or greater | 5,939 (54%) | 1,540 (50%) |
| Missing | 901 | 489 |
| Region | | |
| Northeast | 2,216 (19%) | 679 (19%) |
| Midwest | 2,383 (20%) | 810 (23%) |
| South | 4,790 (40%) | 1,290 (36%) |
| West | 2,522 (21%) | 766 (22%) |
| Missing | 13 | 9 |
| Hospitalization status at initial COVID-19 illness | | |
| Not hospitalized | 10,612 (89%) | 3,305 (93%) |
| Hospitalized | 1,304 (11%) | 245 (6.9%) |
| Missing | 8 | 4 |
| Comorbid conditions | | |
| Diabetes | 561 (4.7%) | 119 (3.4%) |
| Cardiovascular disease/congestive heart failure | 287 (2.4%) | 68 (1.9%) |
| Hypertension | 1,930 (16%) | 492 (14%) |
| Chronic kidney disease | 97 (0.8%) | 32 (0.9%) |
| Cancer | 306 (2.6%) | 71 (2.0%) |
| Asthma/reactive airway disease/chronic lung disease | 2,077 (17%) | 485 (14%) |

**Supplementary Table 2** Comparison of baseline demographic and clinical characteristics among participants with SARS-CoV-2 infection who agreed and declined to be contacted for future studies[1,2,3,4]

| | Agreed to be Contacted for Future Studies (n = 11,924) | Declined to be Contacted for Future Studies (n = 3,554) |
|---|---|---|
| Overweight/obese | 3,670 (31%) | 969 (27%) |
| Autoimmune disorder | 1,203 (10%) | 297 (8.4%) |
| Stroke | 93 (0.8%) | 14 (0.4%) |
| Depression/anxiety/other mental health condition | 4,262 (36%) | 1,171 (33%) |
| Missing | 1 | 0 |
| Long COVID status at survey completion | | |
| Has long COVID | 7,698 (65%) | 2,069 (58%) |
| Does not have long COVID | 693 (5.8%) | 421 (12%) |
| Cannot be determined[4] | 3,533 (30%) | 1,064 (30%) |

[1]Missing data were due to invalid data, “don’t know” responses, “refuse to answer” responses, and missing responses. Percentages do not add up to 100% due to missing data. The number of missing data varies due to participants dropping out of the survey at different sections.

[2]Baseline data in participants with a history of SARS-CoV-2 infection is as of October 20, 2022.

[3]Limited to participants with a defined long COVID status. Long COVID status was determined using the WHO definition. 320 participants without a defined long COVID status were excluded: 283 provided an initial SARS-CoV-2 infection date before the first confirmed positive test in the United States (January 20, 2020), 28 provided an invalid SARS-CoV-2 infection date, nine did not provide a SARS-CoV-2 infection date nor report experiencing symptoms of COVID-19.

[4]Long COVID status could not be determined because fewer than 12 weeks existed between initial SARS-CoV-2 infection and survey completion.
